# Comparison of the analytical and clinical sensitivity of thirty-four rapid antigen tests with prevalent SARS-CoV-2 variants of concern during the COVID-19 pandemic in the UK

**DOI:** 10.1101/2023.07.24.23293072

**Authors:** Rachel L. Byrne, Rachel S. Owen, Ghaith Aljayyoussi, Caitlin Greenland-Bews, Konstantina Kontogianni, Anushri Somasundaran, Dominic Wooding, Christopher T. Williams, LSTM Diagnostics Group, Falcon Steering Group, Margaretha de Vos, Richard Body, Emily R. Adams, Camille Escadafal, Thomas Edwards, Ana I. Cubas-Atienzar

## Abstract

Antigen-detection rapid diagnostic tests (Ag-RDTs) have become a central pillar for the management of COVID-19 worldwide due to their speed and ease of use, and are now being developed for use in other emerging outbreaks. Like other viruses, SARS-CoV-2 is subject to rapid mutation as it spreads, and new Variants of Concern (VOCs) emerge frequently, posing a significant challenge for the reliability of Ag-RDTs designed against the original strain to detect newer, highly mutated variants. It is therefore important that the performance of Ag-RDTs is regularly evaluated, particularly in outbreak scenarios where rapid diagnostics are key to limiting disease spread. Here, we present a comprehensive evaluation of the analytical and clinical sensitivity of 34 commercially available Ag-RDTs with five VOCs, all of which were highly prevalent in the UK at various times between 2019 and 2023. This evaluation highlights the importance of regular evaluation of Ag-RDT performance, with several Ag-RDTs demonstrating reduced performance with some VOCs. This study also highlights the challenges that arise when different organisations lack a consensus approval threshold, causing confusion over Ag-RDT performance and making it difficult to properly evaluate individual tests. We conclude that regular performance evaluation through our proposed pipeline combined with a broad consensus approval threshold across global organisations is essential to maintaining the effectiveness of Ag-RDTs as a disease management tool during outbreaks.

**Importance:** Antigen-detection rapid diagnostic tests (Ag-RDTs) came to global prominence during the COVID-19 pandemic, where they offered a quick and simple at home diagnostic which could be used to manage disease spread. A major ongoing challenge for the broad use of Ag-RDTs is the speed at which new SARS-CoV-2 variants emerge, each of which has the potential to reduce the performance of the available Ag-RDTs. As Ag-RDTs are explored for use in other viral disease outbreaks, pipelines for the regular evaluation of test performance are essential for ensuring that Ag-RDTs can be employed effectively. Here, we have developed a robust pipeline for the large-scale evaluation of commercially available Ag-RDTs against several major SARS-CoV-2 variants, which can be adapted and applied to other emerging outbreaks to ensure test performance is maintained as a virus evolves.

## Introduction

Antigen-detection rapid diagnostic tests (Ag-RDTs) offer quick, inexpensive, laboratory-independent diagnostics that can be performed at home by lay individuals (1,2). As such they have been a central pillar of control strategies for SARS-CoV-2 and are now the first line diagnostic in many countries (3). Whilst the development and deployment of COVID 19 Ag-RDTs was rapid, the frequent emergence and spread of new variants of concern (VOCs) combined with the global nature of the pandemic response posed a continuous challenge to disease monitoring. The majority of Ag-RDTs were developed and evaluated early in the COVID-19 pandemic, utilising wild type (WT) SARS-CoV-2’s nucleocapsid (N) protein as a target, due to its high abundance within the virion (4). However, further VOCs rapidly emerged, including Alpha, Beta, Gamma and Delta, and by the time of the emergence of the first Omicron VOC in late 2021 the viral genome had accumulated a significant number of mutations (5,6). These mutations included several within the N protein, where the Omicron lineage has 3 unique mutations, making it difficult to predict the performance of Ag-RDTs with emerging VOCs (7).

Initial data on the performance of Ag-RDTs for different COVID-19 VOCs remain contradictory in both analytical and clinical evaluations. Whilst early reports evaluating a small number of brands found comparable sensitivities between WT, Delta and Omicron (B.1.1.529) VOCs (8–11), later studies demonstrated that Ag-RDTs showed loss of sensitivity to Omicron when compared to other VOCs (12), and clinical evaluations of Ag-RDTs against emerging SARS-CoV-2 lineages have reported inconsistent results (12–14).

In order for RDTs to be used to their full potential in managing large scale disease outbreaks, there must be confidence that these tests maintain good clinical specificity even as a pathogen evolves, and regular clinical and analytical evaluation is essential to ensuring that RDTs are performing to the required standards. Given the comparatively recent emergence of large scale RDT testing in the context of infectious disease, relatively few evaluations of these devices on a large scale over time have been conducted. Here, we present a robust pipeline for the evaluation of commercially available RDTs against an evolving pathogen, SARS-CoV-2, using the most prevalent VOCs circulating in the United Kingdom between 2019-2023.

## Results

### Analytical sensitivity using cultured SARS-CoV-2 virus

For Omicron sub lineage BA.5, all of the 31 Ag-RDTs evaluated had an LOD ≤ 5.0 × 10^2^ pfu/mL, fulfilling the criteria set by the British Department of Health and Social Care (DHSC) (Fig. 1) and all except two brands (RespiStrip and GeneFinder) had an LOD of ≤ 1.0 × 10^6^ RNA copies/mL thus fulfilling the WHO Target Product Profile (TPP) recommendations for SARS-CoV-2 Ag-RDTs (15). Whereas for Omicron BA.1, only 23 of the 34 Ag-RDTs evaluated had an analytical LOD of ≤ 5.0 × 10^2^ pfu/mL and 32 out of 34 (including Biocredit, Core, Covios, Hotgen, Innova, LumiraDx, PerkinElmer and SureStatus, that all fell below the DHSC recommendations) had an LOD of ≤ 1.0 × 10^6^ RNA copies/mL. The more sensitive Ag-RDTs for detecting Omicron BA.1 were AllTest, Bioperfectus, Flowflex, Fortress, Joysbio, Nadal, Onsite, RightSign, Roche, StrongStep, Standard Q, Tingsun and Wondfo (Fig. 1) with an LOD ≤ 2.5 x 10^2^ pfu/mL and 4.4 x 10^4^ RNA copies/mL. For both Omicron BA.1 and BA.5 the Ag-RDT brand with the lowest sensitivity was RespiStrip with an LOD of 5.0 x 10^4^ pfu/mL and 9.2 x 10^6^ RNA copies/mL (BA.1) and LOD of 1.0 x 10^2^ pfu/mL and 3.5 x 10^6^ RNA copies/mL (BA.5) respectively.

**FIG 1.**
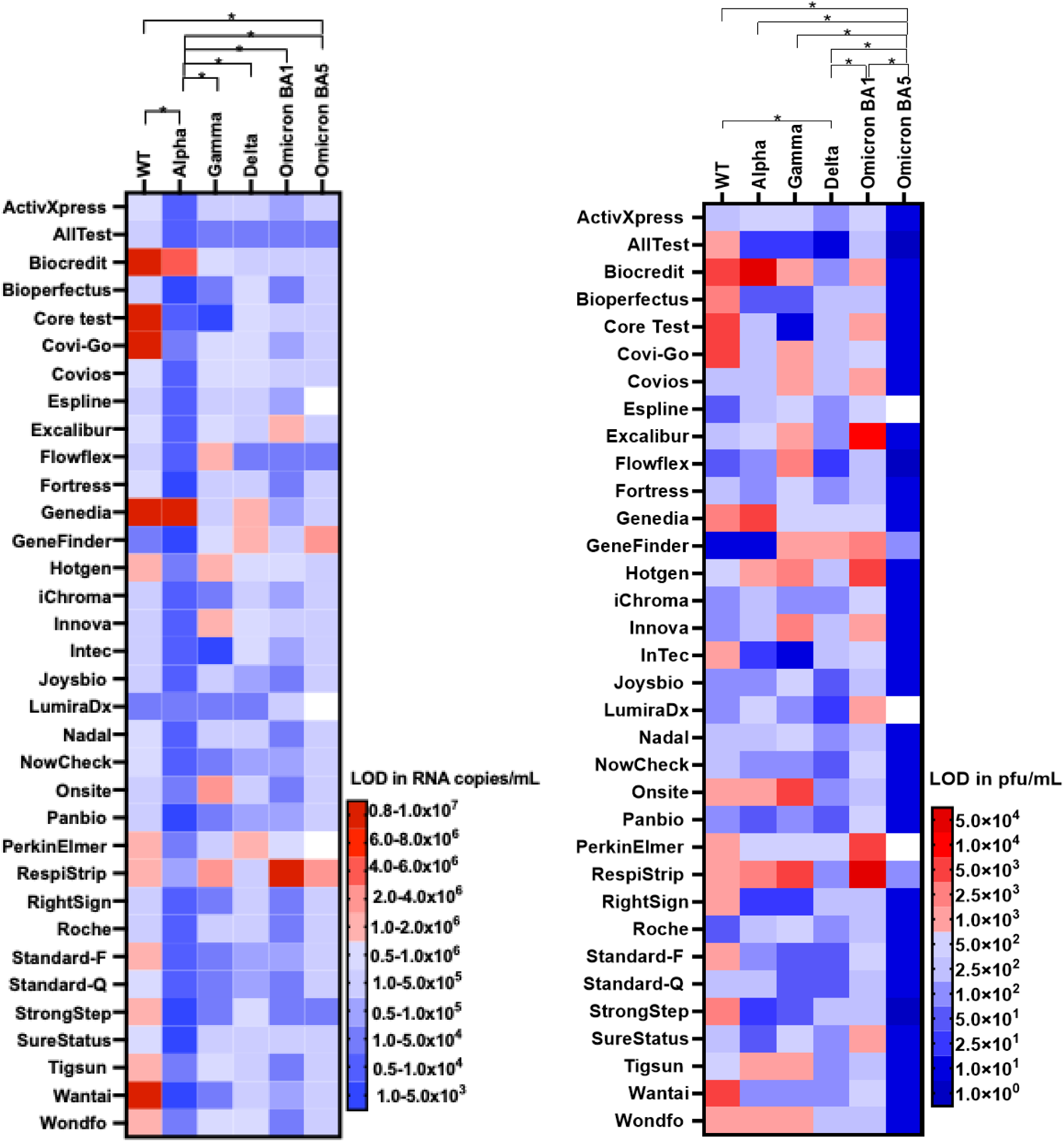
Heatmap comparing the LODs of 34 Ag-RDT using the Ancestral (WT), Alpha (B.1.1.7), Gamma (P.1), Delta (B.1.617.2) Omicron (BA.1) and Omicron (BA.5) variants on pfu/mL (right) and RNA copies/mL (left). Data of the Ancestral, Alpha and Gamma have been taken from our previously published work (16). Blue colours indicate LODs fulfilling the DHSC (for pfu/mL) and WHO criteria (for RNA copies/mL). * = p ≤ 0.05 between VOCs.

For the Delta VOC, 33 out of the 34 Ag-RDTs evaluated had an LOD of ≤ 5.0 × 10^2^ pfu/mL and 31 of the 34 reported an LOD ≤ 1.0 × 10^6^ RNA copies/mL (Fig. 1), with Genefinder falling to meet either the DHSC or WHO recommended LOD.

For the Alpha VOC, 7 of the 34 Ag-RDTs evaluated (Biocredit, Genedia, Hotgen, Onsite, RespiStrip, Tigsun and Wondfo) had an analytical LOD of > 5.0 × 10^2^ pfu/mL thus failing to meet the DHSC minimum requirements, with Biocredit and Genedia also falling below the WHO requirement.

For the Gamma VOC, 5 Ag-RDT brands (Flowflex, Hotgen, Innova, Onsite and RespiStrip) failed to meet either the DHSC or WHO requirements with a further 7 Ag-RDT brands falling below the DHSC recommendations. The Ag-RDTs with the greatest sensitivity for a sample positive for Gamma VOC where AllTest, Core Test, InTec, Standard-F, Standard-Q, StrongStep and Surestatus (Fig. 1).

For the WT, the target for which all Ag-RDT brands were originally developed, only 19 and 22 met the DHSC and WHO requirements, respectively.

When comparing only the pfu/mL, we found that tests had significantly higher LODs with Omicron BA.1 compared to Delta (p = 0.000) and significantly lower LODs with Omicron BA.5 compared to all other VOCs tested (p = 0.001). When comparing RNA copies/mL, the Ag-RDTs detected more sensitively Alpha (p = 0.000) than the other VOCs (p = 0.000).

### Retrospective samples: SARS-CoV-2 Ag-RDT clinical sensitivity

The clinical sensitivity of five Ag-RDTs brands (Covios, Flowflex, Hotgen, Onsite and SureStatus) was evaluated utilising SARS-CoV-2 Alpha (n = 30), Delta (n = 56) and Omicron (n = 49) positive VTM swabs stored at −80 °C, as previously described. Statistically higher viral loads, determined by RT-qPCR, were recorded among individuals positive for Alpha and Omicron infection compared to Delta (p = 0.001 and p = 0.009 respectively) measured by Kruskal–Wallis (Fig. 2).

**FIG 2:**
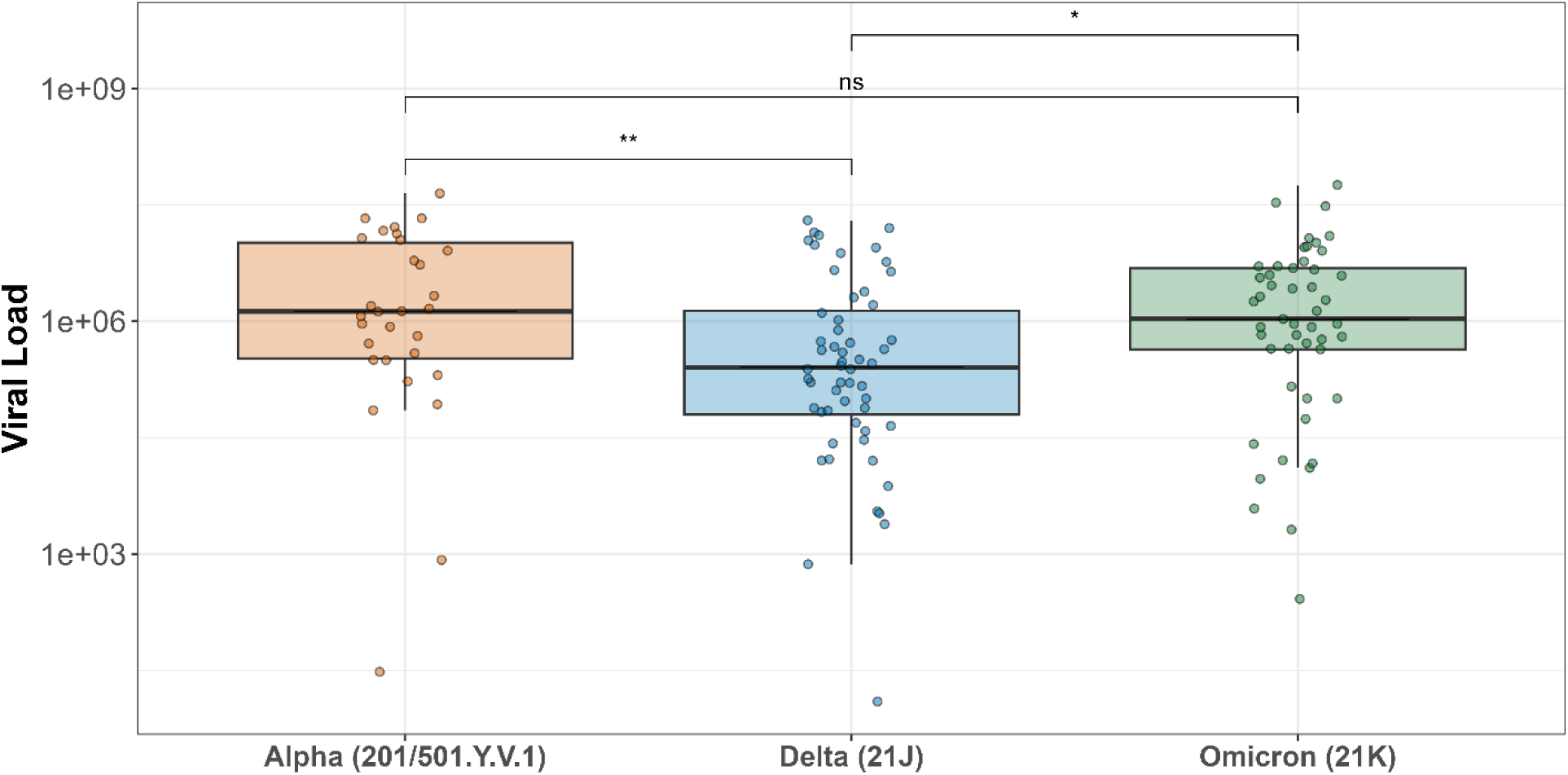
Boxplot of the SARS-CoV-2 viral load (RNA copies/mL) distribution of RT-qPCR NP swabs collected from participants recruited to FALCON between January 2021 and March 2022. The whiskers show the 95 % confidence intervals and the horizontal line the median. Asterisks indicate statistical significance between different VOCs (ns = non-significant, * = p ≤ 0.05, ** = p ≤ 0.01).

We determined the 50 % and 95 % LODs with Alpha, Delta, and Omicron SARS-CoV-2 positive swab samples for five Ag-RDT brands based on a logistic regression model (Table 1, Fig. 3). Overall, the lowest LOD for the Alpha VOC was recorded with Flowflex Ag-RDT (50 % LOD 1.58 x 10^4^ RNA copies/mL and 95 % LOD 2.14 x 10^4^ RNA copies/mL), for Delta variant with Onsite Ag-RDT (50 % LOD 3.31 x 10^1^ RNA copies/mL and 95 % LOD 3.80 x 10^4^ RNA copies/mL) and for Omicron with SureStatus Ag-RDT (50 % LOD 1.78 x 10^3^ RNA copies/mL and 95 % LOD 7.41 x 10^4^ RNA copies/mL), which were all statistically similar to the true analytical LOD reported. The Delta VOC exhibited the greatest variability between predicted LODs from different Ag-RDTs (Table 1) whereas for the true analytical LOD the Alpha VOC showed the greatest variability (Fig. 1).

**TABLE 1:**
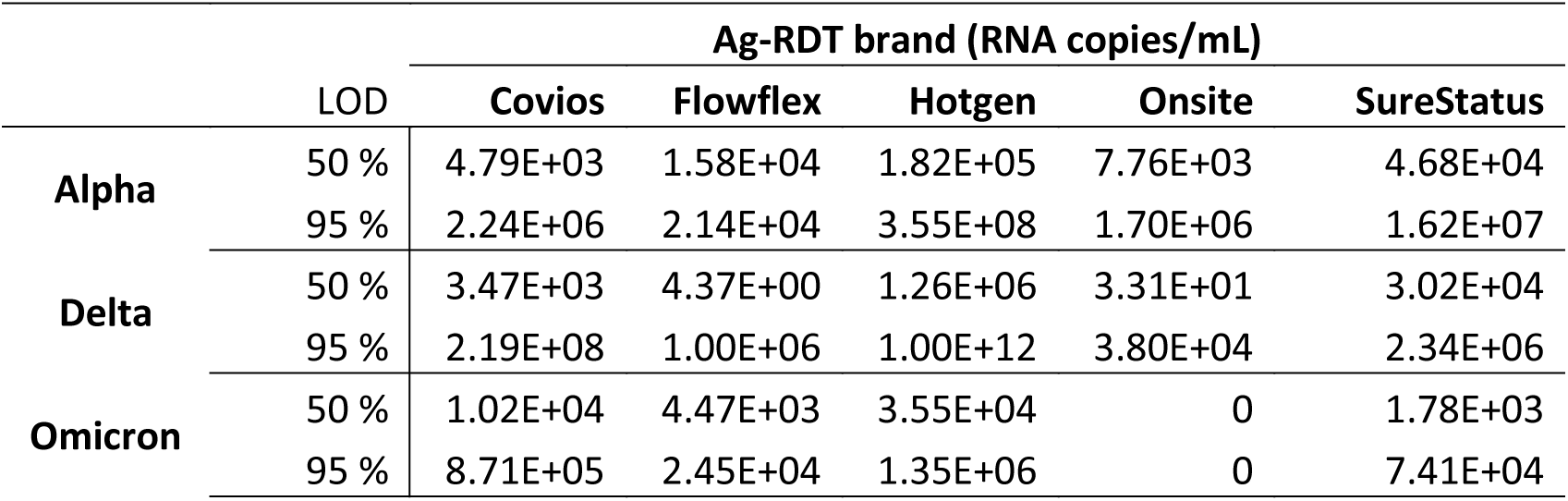
The 50 % and 95 % limit of detection (LOD) (RNA copies/mL) for five Ag-RDT brands (Covios, Flowflex, Hotgen, Onsite and SureStatus) from 122 clinical NP samples positive for Alpha, Delta and Omicron SARS-CoV-2 VOCs.

|  | LOD | Ag-RDT brand (RNA copies/mL) |  |  |  |  |
| --- | --- | --- | --- | --- | --- | --- |
|  |  | Covios | Flowflex | Hotgen | Onsite | SureStatus |
| Alpha | 50 % | 4.79E+03 | 1.58E+04 | 1.82E+05 | 7.76E+03 | 4.68E+04 |
|  | 95 % | 2.24E+06 | 2.14E+04 | 3.55E+08 | 1.70E+06 | 1.62E+07 |
| Delta | 50 % | 3.47E+03 | 4.37E+00 | 1.26E+06 | 3.31E+01 | 3.02E+04 |
|  | 95 % | 2.19E+08 | 1.00E+06 | 1.00E+12 | 3.80E+04 | 2.34E+06 |
| Omicron | 50 % | 1.02E+04 | 4.47E+03 | 3.55E+04 | 0 | 1.78E+03 |
|  | 95 % | 8.71E+05 | 2.45E+04 | 1.35E+06 | 0 | 7.41E+04 |

**FIG 3:**
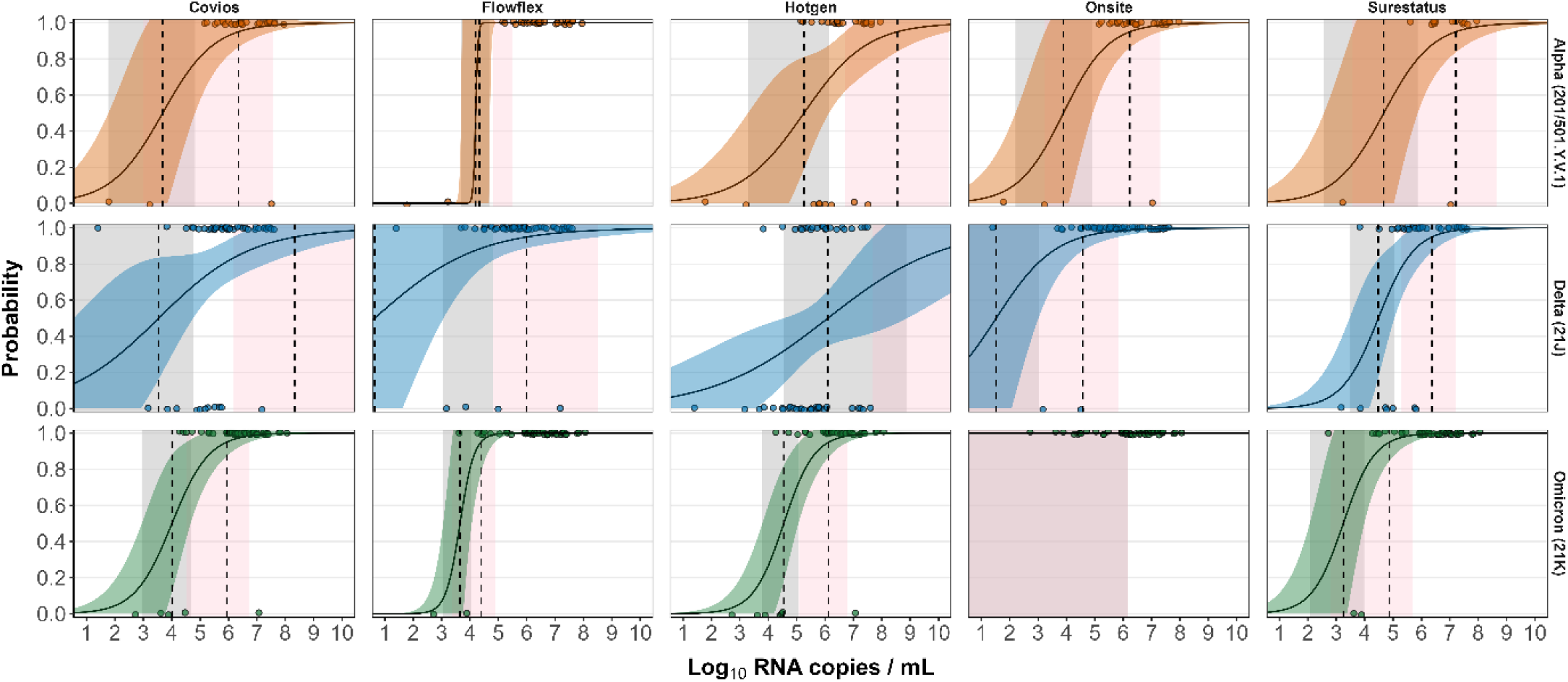
Limit of detection analyses of upper-respiratory samples positive by RT-qPCR for five SARS-CoV-2 Ag-RDT tests (Covios, Hotgen, Onsite, Flowflex and SureStatus) using NP swabs. The log10 RNA copies on the x axis were plotted against a positive (1.0) or negative (0.0) Ag-RDT result on the y axis. Fill curves show logistic regressions of the viral load on the Ag-RDT result; vertical dashed lines indicate log10 RNA copies subjected to the test at which 50 % and 95 % LOD of the samples are expected positive based on the regression results. No significant differences were observed.

The sensitivity of the Ag-RDTs varied from 70 % to 93.3 % (Hotgen and Flowflex) for the Alpha VOC, as shown in Table 2. For Delta VOC, the sensitivity ranged from 77.6 % to 94.5 % (Covios and Onsite) for four of the five Ag-RDTs, with Hotgen reporting a significantly lower sensitivity of 44.2 % than Omicron (p < 0.001) and Alpha (p = 0.026). The observed sensitivities for Omicron were consistent across all brands of Ag-RDTs from 84.4 % and 97.9 % (Hotgen and Onsite). For samples with low Ct values (< 25), the sensitivities were statistically similar for the Covios, Flowflex, Onsite and SureStatus Ag-RDTs but not for the Hotgen Ag-RDT. For the Delta VOC, the sensitivity of the Hotgen Ag-RDT was significantly lower compared to Omicron (p < 0.001) and Alpha (p = 0.011). Whereas samples with high Ct values (Ct > 25) resulted in reduced test sensitivities across all variants, with the greatest sensitivity reported in samples positive with the Omicron VOC. A three-way factorial ANOVA assessing the effects of RDT result (value: positive vs. negative), VOC, and RDT brand, along with their interactions, revealed a significant main effect of RDT result (F(1, 593) = 98.21, p < 0.001, η²p = 0.14), indicating a large effect size (Fig. 4). The effect of VOC was also significant (F(2, 593) = 27.83, p < 0.001, η²p = 0.09), while RDT brand had a small and non-significant effect (F(4, 593) = 1.58, p = 0.18, η²p = 0.01). The interaction between RDT result and VOC was significant (F(2, 593) = 14.20, p < 0.001, η²p = 0.05), suggesting that the effect of Ct value on RDT result differs across VOCs. Other interactions, including the three-way interaction, showed negligible effect sizes (η²p < 0.01) and were not significant.

**TABLE 2:** The comparison of the sensitivity of five antigen rapid diagnostic tests (Ag-RDTs) between the Alpha, Delta and Omicron variants. Calculated sensitivities, sample size (in italics) and 95 % confidence intervals (CI95 %) are given.

| Population | Variant | Covios | Flowflex | Onsite | Hotgen | Surestatus |
| --- | --- | --- | --- | --- | --- | --- |
| **Total cohort** | Alpha | 90%, *30* | 93.3%, *30* | 90%, *30* | 70%, *30* | 86.7%, *15* |
|  | (CI95%) | 73.5-97.9% | 77.9-99.2% | 73.5-97.9% | 50.6-85.3% | 59.5-98.3% |
|  | Delta | 77.6%, *49* | 92.9%, *56* | 94.5%, *55* | 44.2%, *52* | 81.6%, *38* |
|  | (CI95%) | 63.4-88.2% | 82.7-98% | 84.9-98.9% | 30.5-58.7% | 65.7-92.3% |
| **Total cohort** | Omicron | 89.8%, *49* | 95.8%, *48* | 97.9%, *47* | 84.4%, *45* | 95.9%, *49* |
|  | (CI95%) | 77.8-96.6% | 85.7-99.5% | 88.7-99.9% | 70.5-93.5% | 86-99.5% |
|  | All | 85.2%, *128* | 94%, *134* | 94.7%, *132* | 64.6%, *127* | 89.2%, *102* |
|  | (CI95%) | 77.8-90.8% | 88.6-97.4% | 89.4-97.8% | 55.6-72.8% | 81.5-94.5% |
| **≤ Ct 25** | Alpha | 95.8%, *24* | 100%, *24* | 95.8%, *24* | 83.3%, *24* | 92.3%, *13* |
|  | (CI95%) | 78.9-99.9% | 85.8-100% | 78.9-99.9% | 62.6-95.3% | 64-99.8% |
|  | Delta | 81.8%, *33* | 94.6%, *37* | 100%, *36* | 45.5%, *33* | 88%, *25* |
|  | (CI95%) | 64.5-93% | 81.8-99.3% | 90.3-100% | 28.1-63.6% | 68.8-97.5% |
| **≤ Ct 25** | Omicron | 97.6%, *42* | 100%, *41* | 100%, *40* | 97.4%, *38* | 100%, *42* |
|  | (CI95%) | 87.4-99.9% | 91.4-100% | 91.2-100% | 86.2-99.9% | 91.6-100% |
|  | All | 91.9%, *99* | 98%, *102* | 99%, *100* | 75.8%, *95* | 95%, *80* |
|  | (CI95%) | 84.7-96.4% | 93.1-99.8% | 94.6-100% | 65.9-84% | 87.7-98.6% |
| **> Ct 25** | Alpha | 66.7%, *6* | 66.7%, *6* | 66.7%, *6* | 16.7%, *6* | 50%, *2* |
|  | (CI95%) | 22.3-95.7% | 22.3-95.7% | 22.3-95.7% | 0.4-64.1% | 1.3-98.7% |
|  | Delta | 68.8%, *16* | 89.5%, *19* | 84.2%, *19* | 42.1%, *19* | 69.2%, *13* |
|  | (CI95%) | 41.3-89% | 66.9-98.7% | 60.4-96.6% | 20.3-66.5% | 38.6-90.9% |
| **> Ct 25** | Omicron | 42.9%, *7* | 71.4%, *7* | 85.7%, *7* | 14.3%, *7* | 71.4%, *7* |
|  | (CI95%) | 9.9-81.6% | 29-96.3% | 42.1-99.6% | 0.4-57.9% | 29-96.3% |
|  | All | 62.1%, *29* | 81.2%, *32* | 81.2%, *32* | 31.2%, *32* | 68.2%, *22* |
|  | (CI95%) | 42.3-79.3% | 63.6-92.8% | 63.6-92.8% | 16.1-50% | 45.1-86.1% |

Ct = cycle threshold

**FIG 4:**
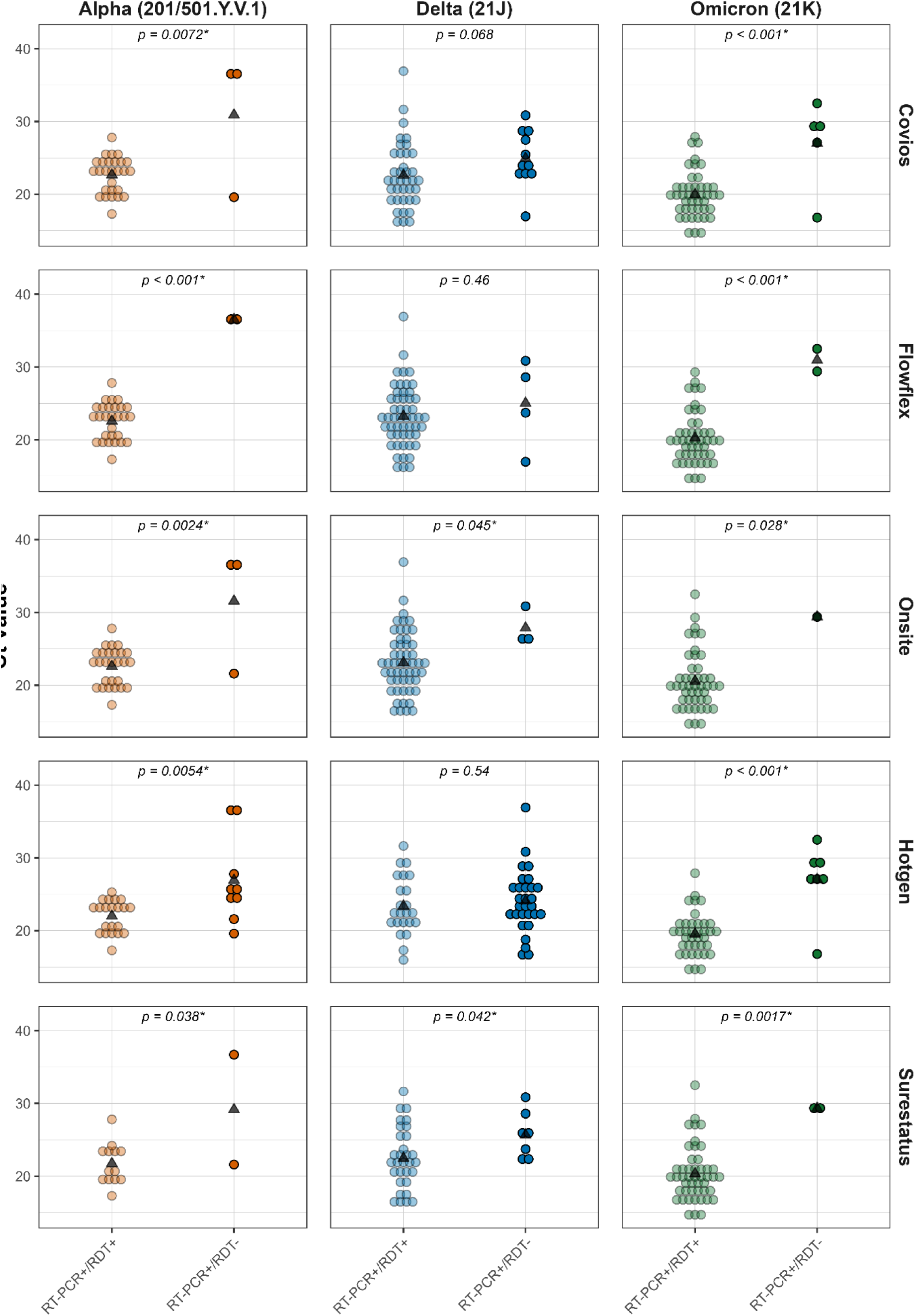
Positive and negative Ag-RDT results according to RT-qPCR cycle threshold (Ct) values. P values indicate the results of Bonferroni-adjusted post-hoc tests performed following three-way factorial ANOVA analysis to compare RDT+ and RDT− within each VOC and RDT brand. Asterisks indicate comparisons where Cohen’s d ≥ 0.5 and p < 0.05, indicating a significant difference between the Genomic Ct values of RT-PCR+/RDT+ samples and RT-PCR+/RDT-samples.

### Prospective samples: SARS-CoV-2 Ag-RDT clinical sensitivity

During the prospective evaluation, 122 participants tested positive by RT-qPCR for SARS-CoV-2. Of them, 32 were the Delta VOC and 90 the Omicron (BA.1) VOC. Samples were immediately tested using the Biocredit (RapiGEN, South Korea) Ag-RDT with 99 yielding a positive Biocredit Ag-RDT result (Sens 81.1 %, CI95 % 73.1-87.6 %). When separated by VOC the sensitivity was lower, but statistically similar, for the Delta VOC 71.9 % (CI 95 % 53.3-86.6 %) compared to the Omicron VOC 84.4 % (CI95 % 75.3-91.2 %). The difference in Ct value between positive and negative samples by Biocredit was significant (Delta p = 0.038, Omicron p = 0.00007) across both VOCs. Additionally, the intensity of the test band of positive Ag-RDTs was recorded for 119 samples (three excluded due to line intensity not recorded) with no significance obtained between the two VOCs (Delta and Omicron BA.1).

## Discussion

We present a comprehensive assessment of the analytical sensitivity of 34 commercially available COVID-19 Ag-RDTs for the detection of the Omicron (BA.1 and BA.5), Alpha, Gamma and Delta VOCs and WT, the major variants circulating in the UK during the initial five years following the emergence of the pandemic(5). The rapid emergence of new VOCs throughout the course of the Covid-19 pandemic posed a significant challenge for reliable rapid diagnostic testing(13). New VOCs often contained mutations in the N gene, increasing the likelihood that RDT analytical sensitivity could be compromised. Indeed, studies have identified mutations in the N gene which affected the sensitivity of Ag-RDTs, including the T135I mutation in the Alpha VOC (17), and the A376T and M241I mutations (18), thus it is important to maintain regular assessment of Ag-RDT performance over time.

Analytically, the majority of Ag-RDTs successfully met the WHO criteria outlined in the TPP for SARS-COV-2 Ag-RDTs when detecting the Omicron viral lineages (94.1 %, BA.1 and 93.5 %, BA.5)(15). When analysing RNA copies/mL, as outlined in the WHO recommendations, the LOD of Omicron VOC BA.5 was statistically lower than the WT and the Alpha VOC but similar to the Gamma, Delta and Omicron BA.1 VOCs. However, when comparing the PFU/mL, recommended by the UK DHSC, tests have significantly lower LODs with Omicron BA.5 than all other VOCs and WT.

The observed discrepancies between RNA copies/mL and PFU/mL are likely attributable to difference in the virus’ ability to form plaques and varying ratios of infectious particles to RNA copies present (19). Whilst the PFU/mL measures infectious virus present, RNA copies/mL encompasses all SARS-CoV-2 RNA present including non-infectious or dead virus. For Ag-RDT evaluation, RNA copies/mL is recommended by the WHO and the UK DHSC recommends PFU/mL. It is counterintuitive for organisations to utilise different units of measurements for comparison of Ag-RDTS and a consensus would allow more appropriate evaluation, something that is essential for effectively managing responses to new disease outbreaks.

For 5 of the 34 Ag-RDT brands we present clinical accuracy data utilising clinical respiratory samples positive for SARS-CoV-2 Alpha, Delta and Omicron (BA.1) VOCs. Hotgen consistently performed poorer than other Ag-RDTs, typically falling below the WHO TPP guidance with Onsite the best performing brand overall. For all brands, the clinical sensitivity values obtained with retrospective samples were the highest for Omicron samples. This is in line with other published works comparing Omicron BA.1 to other non-Omicron VOCs (20). A similar trend was observed when using only prospective samples collected when the UK was experiencing the Delta and Omicron waves of infection. A lower sensitivity was recorded for samples positive for the Delta VOC compared to those positive with an Omicron VOC infection on the Biocredit Ag-RDT, as shown in cultured samples by Stanley *et al.,* 2022 (21). These differences in sensitivity of Ag-RDTs between VOCs, which are independent of the temporal emergence of each variant, highlight the importance of continual and thorough assessment of RDT performance as new lineages of a pathogen emerge.

For all brands in the retrospective cohort the predicted clinical LODs were statistically similar to the analytical LODs obtained with spiked laboratory samples. Clinical evaluation is expensive, and often surplus samples are not available for comprehensive testing, hindering large scale assessment of Ag-RDT performance. The data presented here demonstrates the efficacy of analytical samples, indicating that analytical evaluation is a robust alternative for evaluating Ag-RDT performance in the absence of abundant clinical samples.

This study has several strengths; we have carried out an extensive evaluation of the analytical sensitivity of 34 commercially available Ag-RDT brands. This list is inclusive of most WHO-EUL recommended tests and five awaiting approvals at the time of the study thus of high global high public health relevance. Additionally, we included both viral isolates and clinical specimens to evaluate the Ag-RDT sensitivity. The clinical specimens used in this study are attributed to three different lineages; Alpha, Delta and Omicron, which were all at some point a dominant VOC, providing a comprehensive evaluation of assay performance across a significant temporal and mutational timeframe.

Limitations of this study include the use of retrospective frozen specimens instead of fresh swabs as recommended by most Ag-RDT manufacturers. However, prospective clinical evaluation studies rarely include multiple VOCs as their prevalence depends on their time period, and prospective evaluation of multiple RDT brands simultaneously is complicated by the need for a single swab per test. To correct for the potential degradation of RNA after a freeze-thaw cycle, viral RNA was re-tested by RT-PCR at the time of Ag-RDT evaluation and these values were used for comparison. Another limitation of this study is that we did not repeat the LOD experiments for the WT, Alpha and Gamma viral isolates and used the data from previous studies performed in our laboratory with the same protocol and compared it to the newly produced data for the Delta and Omicron VOC isolates.

To conclude, we present a robust pipeline for the large scale evaluation of commercially available Ag-RDTs against multiple SARS-CoV-2 VOCs, which can be easily adapted to assess further variants or for different disease outbreaks. We report similar if not superior LODs for Omicron compared to other non-Omicron VOCs across a wide range of commercially available RDTs. However, we also report decreased detection of the Delta VOC in both analytical and clinical samples, highlighting the need for continuous assessment of Ag-RDTs especially those recommended for at home testing. We additionally report inconsistencies between product fulfilment criteria for WHO and UK DHSC, where several tests show significantly different performance depending on the guidelines, highlighting the importance of standardised evaluation criteria for Ag-RDTs, particularly during a global response.

## Materials and Methods

### Evaluated Ag-RDTs

Thirty-four Ag-RDT brands were evaluated in this study; all were lateral flow assays (LFA), 31 using colorimetric gold nanoparticle detection, two fluorescence and one based on microfluidic immunofluorescence technology (Table 3). The selection of the Ag-RDT resulted from an expression of interest launched by FIND (www.finddx.org) and a scoring process based on defined criteria. This list includes eight Ag-RDTs on the WHO Emergency Use Listing (WHO-EUL) and six tests that are on the waiting list for WHO-EUL approval (22). Analytical testing was performed on all Ag-RDT brands (Table 3) and a small subset of these were further used for the clinical evaluation on retrospective samples based on brands that showed the best results on clinical diagnostic evaluations under the FIND programme (Covios, Hotgen, Onsite, SureStatus) and widely used in the UK for mass testing (Flowflex). Results on prospectively collected samples are only provided with Biocredit.

**TABLE 3:** An overview of the RDT brands used in the study for analytical performance.

| RDT brand | Test/Company/Country | Target Ag | Principle | Approval |
| --- | --- | --- | --- | --- |
| ActiveXpress | ActivXpress+ COVID-19 Ag Complete Kit/Edinburgh Genetics Ltd./UK | N | G | CE |
| AllTest | SARS-CoV-2 Antigen Rapid Test/Hangzhou AllTest Biotech Ltd./China | N | G | CE |
| Biocredit | Biocredit COVID-19 Ag/Rapidgen Inc./Rep. Korea | N | G | CE/ WHO EUL |
| Bioperfectus | SARS-CoV-2 Ag Rapid Test/ Jiangsu Bioperfectus Tech. Ltd./China | N | G | CE |
| Core | COVID-19 Ag Test/Core Technology Ltd./China | N | G | CE |
| Covi-Go | Covigo/Mologic Ltd./UK | N | G | CE/ UA |
| Covios | Covios COVID-19 Ag Test device/ Mologic Ltd./UK | N | G | CE |
| Espline | ESPLINE SARS-CoV-2/Fujirebio Diagnostics Inc./Japan | N | G | CE |
| Excalibur | Rapid SARS-CoV-2 Antigen test card/ Excalibur Healthcare Services/UK | N | G | CE |
| Flowflex | Flowflex SARS-CoV-2 Ag Rapid Test/Acon Biotech, Ltd./China | N | G | CE/ WHO EUL |
| Fortress | CoV Ag Nasal Swab Rapid test/Zhejiang Orient Gene Biotech/ China | N | G | CE |
| Genedia | GENEDIA W COVID-19 Ag/ Green Cross Medical Sciences/Rep. Korea | N | G | CE |
| GeneFinder | COVID-19 rapid test GeneFinder/OSANG Health Care/ South Korea | N | G | CE/UA |
| Hotgen | 2019-nCoV Antigen test/ Beijin Hotgen Biotech Ltd./China | N | G | CE |
| iChroma | iChroma COVID-19 Ag Test/ Boditech Medical Inc./Rep. Korea | N | F | CE |
| Innova | Innova SARS-CoV-2 Antigen Rapid/ Innova Medical Group Ltd./UK | N | G | CE/UA |
| Intec | Rapid SARS-CoV-2 Antigen test/Intec Products Inc./China | N | G | CE |
| Joysbio | SARS-CoV-2 Antigen Rapid Test Kit/ Joysbio Biotechnology Ltd./China | N | G | CE/UA |
| LumiraDx | LumiraDx SARS-CoV-2 antigen test/ Lumira Dx Ltd./US | N | M | CE/ WHO EUL |
| Nadal | Nadal COVID-19 Ag Test/Nal von minden GmbH/Germany | N | G | CE |
| NowCheck | NowCheck COVID-19 Ag test/ Bionote Inc./ Rep. Korea | N | G | CE |
| Onsite | Onsite COVID-19 Ag Rapid Test/CTKBiotech Inc./USA | N | G | CE/ WHO EUL |
| PanBio | Panbio COVID-19 Ag Rapid Test/Abbott Rapid Diagnostics/Rep. Korea | N | G | CE/WHO EUL |
| PerkinElmer | PerkinElmer COVID-19 Antigen Test/PerkinElmer/ Switzerland | N | G | CE/UA |
| RespiStrip | Respi-Strip COVID-19 Ag/Coris Bioconcept/Belgium | N | G | CE |
| RighSign | COVID-19 Ag Rapid Test Cassette/Hangzhou Biotets Biotech Ltd./China | N | G | CE |
| Roche | SARS-CoV-2 Rapid Ag Test/ Roche Diagnostics/Switzerland | N | G | CE |
| Standard-F | Standard F COVID-19 Ag FIA., SD Biosensor Inc./Rep. Korea | N | F | CE |
| Standard-Q | Standard Q COVID-19, SD Biosensor Inc./Rep. Korea | N | G | CE/WHO EUL |
| StrongStep | StrongStep SARS-CoV-2 Ag Rapid Test/Nanjing Liming Bio-Products/US | N | G | CE |
| SureStatus | Sure-Status COVID-19 Antigen Card Test, Premier Medical Corp./ India | N | G | WHO EUL |
| Tigsun | Tingsun COVID-19 Ag Rapid test/ Beijin Tigsun Diagnostics Ltd./China | N | G | CE |
| Wantai | Rapid SARS-CoV-2 Antigen test/ Wantai Biological Pharmacy Ltd./China | N | G | CE |
| Wondfo | Wondfo 2019-nCoV Antigen Test/ Guangzhou Wondfo Biotech/China | N | G | CE/WHO EUL |

RDT: Rapid diagnostic test. Target N: nucleoprotein; Principle G: LFA using gold (colourimetric detection), F: LFA
fluoresce detection, M: microfluidic fluorescent technology. CE: CE making as per European Conformity, WHO EUL:
WHO Emergency Use Listing, UA: under assessment for WHO Emergency Use Listing.

### SARS-CoV-2 viral culture and Ag-RDT limit of detection

The SARS-CoV-2 isolates were grown in Vero E6 cells (C1008; African green monkey kidney cells) and maintained in culture media (Dulbecco’s modified eagle membrane (DMEM) with 2% fetal bovine serum and 0.05 mg/mL gentamycin) as previously described (23,24). The isolates for Alpha (Genbank accession number: MW980115), Delta (SARS-CoV-2/human/GBR/Liv_273/2021), Gamma (hCoV-19/Japan/TY7-503/2021), Omicron (BA.1) (SARS-CoV-2/human/GBR/Liv_1326/2021) and Omicron (BA.5)(SARS-CoV-2/South Africa/CERI-KRISP-K040013/2022) were used evaluate the analytical limit of detection (LOD) of the 34 Ag-RDTs using live virus.

Plaque forming units per millilitre (pfu/mL) of the viral stocks were counted using viral plaque assay as previously described (16) and ten-fold serial dilutions of the viral stock were made starting from 1.0 x 10^6^ pfu/mL using DMEM as a diluent. Two-fold dilutions were made below the ten-fold LOD dilution to determine the LOD. The LOD was defined as the lowest dilution at which all three replicates were positive by Ag-RDT. The LODs for WT, Alpha and Gamma VOCs, obtained as part of our previous work utilising the same protocol (16,24), were used here for practicality to compare to the Delta and Omicron lineages LODs.

### Retrospective and prospective clinical samples

Clinical samples were collected as part of the ‘Facilitating Accelerated Clinical Evaluation of Novel Diagnostic Tests for COVID-19’ (FALCON) study (25). Ethical approval was obtained from the National Research Ethics Service and the Health Research Authority (IRAS ID:28422, clinical trial ID: NCT04408170). Nasopharyngeal (NP) swabs in vital transport media (VTM) were collected from consenting symptomatic adults attending the community drive-through COVID-19 test center located in Liverpool John Lennon Airport, UK between January 2021 and March 2022. Clinical specimens were transported to the Liverpool School of Tropical Medicine (LSTM) biosafety level 3 (BSL3) laboratories in insulated UN7737 transport bags and aliquoted and stored at −80°C until further testing.

Prospective clinical samples were collected as part of a diagnostic evaluation of the Biocredit Ag-RDT (Table 3) (26). Participants were enrolled from December 2021 and March 2022 coinciding with the emergence of Omicron. NP swabs in VTM were collected for RT-qPCR followed by another NP swab in the alternate nostril for the BioCredit Ag-RDT evaluation. Specimens were transported to the LSTM BSL3 laboratories as described above and, in this case, processed immediately for Ag-RDT testing.

Samples were confirmed SARS-CoV-2 RNA-positive using the TaqPath™ COVID19 CE-IVD RT-PCR Kit (Thermo Fisher Scientific, USA). Based upon epidemiological data in the UK at the time of enrolment and S gene amplification in the PCR assay, consecutive SARS-CoV-2 positive RT-PCR samples were selected as presumed Alpha if collected between January and March 2021, presumed Delta if collected between June and August 2021, and presumed Omicron if collected between December 2021 and March 2022. The variant type was later confirmed by whole genome sequencing. A panel of ten SARS-CoV-2 RNA-negative VTM samples were also included as negative controls.

### Ag-RDTs testing protocol

All Ag-RDTs were performed as specified by the test specific instructions for use (IFU). For the determination of the LOD using live virus, a specific volume of the serial dilutions was added directly to the extraction buffers at a 1:10 ratio as previously described (23). For the clinical samples, VTM was mixed by pipetting at 1:1 ratio with the extraction buffer of the Ag-RDTs. The dilution factor introduced by the swabs diluted in buffer was accounted for when calculating the viral copy numbers of the tested swab samples. A negative control of only VTM was incorporated to account for any non-specific reaction as previously reported for some Ag-RDT brands when using VTM (23). Results were read by two operators, blinded to each other and if a discrepant result occurred, a third operator acted as a tiebreaker. The visual read out of the Ag-RDT test band was scored on a quantitative scale from 1 (weak positive) - 10 (strong positive). Ag-RDT results were classified as invalid when the control line was absent.

### Quantification of viral loads

For quantification of the RNA copy numbers per mL (RNA copies/mL) viral RNA was extracted using QIAmp Viral RNA mini kit (Qiagen, Germany) according to the manufacturer’s instructions. The RNA copies/mL were established using the COVID-19 Genesig RT-qPCR kit (PrimerDesign, UK). RT-qPCR testing was carried out using the Rotor-Gene Q (Qiagen, Germany), with a ten-fold serial dilution of quantified in vitro-transcribed RNA incorporated for each PCR run (27). A total of five replicates were tested for each standard curve point, and extracted RNA from each culture dilution was tested in triplicate. The RNA copies/mL for samples was then calculated from the mean Ct value of these replicates.

### Whole genome sequencing

Clinical samples underwent whole genome sequencing to confirm the SARS-CoV-2 variant. Sequencing was performed using the ARTIC V3 (LoCost) (28)sequencing protocol on the MinION R.9.4.1 flow cell (Oxford Nanopore Technology, UK). RT-PCR was performed with a two-step PCR initially the Arctic RT PCR 5X LunaScript® RT SuperMix (New England Biolabs, USA) with 8 μL of RNA sample, and a thermal cycling profile of 2 minutes at 25 °C followed by 10 minutes at 55 °C and then one minute at 95 °C. This was then followed by the Q5^®^ Hot Start High-Fidelity 2X Master Mix (New England Biolabs, USA) using 10 μM of the ARTIC V4.1 primer pools (Integrated DNA Technologies, USA), and a thermal cycling profile of 30 second at 98 °C for heat inactivation, followed by 25 cycles of a 15 second denaturation at 98 °C and a five-minute annealing/extension at 65 °C. Library preparation was carried out using the Ligation Sequencing Kit (SQK-LSK109) and Native Barcoding Expansion Kits (EXP-NBD104 and EXP-NBD114) (Oxford Nanopore Technologies). Basecalling was carried out via MinKnow (v4.2.8), with demultiplexing and read filtering using Guppy (v5.0.7.). The ARTIC pipeline was then used to assemble a consensus genome, BAM files, and variant calling file with *--normalise 200 --threads 4.* Variant calling was carried out using EPI2ME Desktop Agent v3.3.0 with the ARTIC+NextStrain analysis pipeline.

## Statistical analysis

Statistical analyses were performed using SPSS V.28.0, Epi Info V3.01 and R scripts. Binomial confidence intervals for sensitivities and specificities were computed using the Wilson score interval. Differences in the analytical LODs of VOCs were compared using Kruskal Wallis with Bonferroni correction for multiple tests. To further analyse analytical sensitivities, we used logistic regression, with RNA copy number as the independent and test outcomes as the dependent variable, yielding detection probabilities for each viral load level. A three-way factorial ANOVA was performed on log-transformed Genomic Ct values to assess the effects of RDT result (value: positive vs. negative), VOC, and RDT brand, along with their interactions. Bonferroni-adjusted post-hoc tests were performed to compare RDT+ and RDT− within each VOC and RDT brand.

## Data Availability

All data produced in the present study are available upon reasonable request to the authors.

## Acknowledgements.

This work was funded as part of FIND’s work as co-convener of the diagnostics pillar of the Access to COVID-19 Tools (ACT) Accelerator, including support from Unitaid [grant number: 2019-32-FIND MDR], the governments of the Netherlands [grant number: MINBUZA-2020.961444] and from UK Department for International Development [grant number 300341-102]. The FALCON study was funded by the National Institute for Health Research, Asthma UK and the British Lung Foundation. This study received funding by a UKRI-Medical Research Council (MR/R015678/1) MRC/CASE scholarship to RLB and CGB. The funders had no role in study design, data collection and interpretation, or the decision to submit the work for publication. We thank all participants who volunteered to take part in the study. In the United Kingdom, special thanks go to the NIHR Clinical Research Network (CRN) for their support with the recruitment, specially to Sue Dowling, to the LSTM Diagnostic group—Thomas Edwards, Christopher T Williams, Kate Buist, Karina Clerking, Lorna Finch, Helen Savage, Caitlin R Thompson and Rachel Watkins for recruitment, sample collection, and processing, and to the CONDOR steering group—A. Joy Allen, Julian Braybrook, Peter Buckle, Paul Dark, Kerrie Davis, Adam Gordon, Colette Inkson, Dan Lasserson, Clare Lendrem, Andrew Lewington, Mary Logan, Massimo Micocci, Brian Nicholson, Rafael Perera-Salazar, Graham Prestwich, D. Ashley Price, Charles Reynard, John Simpson, Valerie Tate, Philip Turner and Mark Wilcox—for oversight of the trial in the United Kingdom. We would like to thank the facilitators of the following SARS-CoV-2 isolates obtained through BEI Resources: Isolate hCoV-19/South Africa/CERI-KRISP-K040013/2022 (Lineage BA.5; Omicron Variant), NR-56798, deposited by Dr. Alex Sigal.

